# iTreg mediated TGF-Β1 therapy improves functional engraftment of cell therapy in rd1 Retinitis Pigmentosa mouse model

**DOI:** 10.1101/2024.05.16.24307466

**Authors:** K Varsha Mohan, Alaknanda Mishra, Prakriti Sinha, Abaranjitha Muniyasamy, Perumal Nagarajan, Kiran Chaudhary, Pramod Upadhyay

## Abstract

**Purpose:** Retinitis Pigmentosa (RP) is a progressive and hereditary disease that primarily affects the retina, leading to partial or complete vision loss. In addition to the direct impact on vision, the degeneration of the retina in RP also leads to inflammation in the eye, which can further damage the retina and make it difficult to treat the condition with cell therapy. This inflammation led to oxidative stress and cell death, creating an unfavourable environment for the introduction of new cells via cell therapy.

**Methods:** The potential of Transforming Growth Factor-Beta1 (TGF-B1) as an anti-inflammatory agent to treat ocular inflammation was investigated done by administering TGF-B1 intravitreally to the eyes of rd1 mice. However, due to the transient effect of TGF-B1 injection, the in-vitro-induced Treg (iTregs) cells that secrete TGF-B1, were generated and transplanted into the conjunctiva of 4 weeks old rd1 mice to achieve a sustained release of TGF-B1. After administering iTregs, Retinal Neuron-Like Cells (RNLCs) were transplanted into the rd1 mouse retina as a form of cell therapy to improve vision perception.

Flow cytometry was used to estimate the number of Qtracker labelled RNLCs post 30 days of transplantation. The potential of iTregs as an adjunct transplantation with RNLCs to improve cell therapy survival and vision rescue was investigated by conducting Electroretinography and behavioural studies.

**Results:** The study found that ocular inflammation can be reduced by treating with TGF-B1. After 30 days, mice transplanted with iTregs showed a significant increase in the number of transplanted RNLCs that survived compared to the mice who only received RNLCs. In the total fluid of the eye (aqueous plus vitreous), there was a significant increase in the levels of anti-inflammatory cytokines TGF-B1 and IL-10, and some decrease in the levels of pro-inflammatory cytokines Monocyte Chemoattractant Protein-1 (MCP1). The adjunct therapy of iTregs transplantation resulted in improvement in ERG wave functions and vision preservation compared to the group without adjunct iTregs.

**Conclusions:** The administration of TGF-B1-secreting iTregs to the affected eye reduced the inflammatory environment, which enabled transplanted RNLCs to stay longer compared to without TGF-B1. The iTregs mediated sustained anti-inflammatory adjunct therapy can improve the outcome of cell therapy for RP.

## Introduction

Retinitis Pigmentosa (RP) is a heterogeneous group of mutations in over 100 genes that cause progressive photoreceptor loss. The loss of photoreceptors leads to complete or partial vision loss and the current therapeutic measures including vitamin supplementation [1] have failed to have any favourable clinical outcome. Thus, gene therapy [2, 3] and cell-based therapies [4, 5] are being explored for treatment opportunities in RP.

However, RP is further complicated by non-genetic factors [6, 7] such as ocular inflammation [8-10] which exacerbate the disease and limit the potential of cell-based therapies for vision rescue [8, 11-22].

Studies indicate that photoreceptor damage due to the mutation potential potentiates the inflammatory changes [8, 22, 23] in the ocular milieu which is predominantly anti-inflammatory in normal conditions. These changes render the retina hostile to the engraftment of cell-based therapies.

Since the ocular changes are potentiated by the pro-inflammatory changes in the eye, the use of anti-inflammatory molecules as a neutralizing agent has generated scientific interest [8, 17, 24-26]. Transforming Growth Factor-Beta1 (TGF-B1) is a key anti-inflammatory modulator in the eye. The ocular environment is rich in TGF-B1 which counters the damaging functions of Tumor necrosis factor-alpha (TNFα) and microglial activation. Intravitreal TGF-B1 has proved to be beneficial for endotoxin-mediated ocular inflammation [15] and uveitis [19]. Further, in RP the protective and therapeutic potential of TGF-B1 has been demonstrated by ameliorating the neurotoxic inflammatory microglial function by Adeno-Associated Virus (AAV) vector-mediated delivery of TGF-B isoforms to rescue cone neurodegeneration in the mouse models of RP [27].

The Regulatory T cells (Tregs) have also been explored as a stable source of TGF-B1 for managing ocular inflammation. Systemic administration of stimulated-antigen-stimulated Tregs have resulted in the reduction of uveitis [18]. In another study, the localized sub-conjunctival administration of Tregs supported the survival of corneal grafts in rats [28].

We have demonstrated earlier that all mutations of RP induce ocular inflammation and exacerbate retinal loss [12]. Thus, we focused on TGF-B1, which is a key ocular resident anti-inflammatory cytokine [25, 29-31]. It is known to be a balancing factor in the eye and plays a crucial function in maintaining ocular integrity [29]. Thus, we hypothesized that a sustained anti-inflammatory intervention by transplanting TGF-B1 secreting *in-vitro* induced Tregs (iTregs) can potentially reduce the pace of degeneration and create a non-hostile environment for cellular vision rescue in RP.

We transplanted iTregs in the sub-conjunctiva, leading to a sustained release of TGF-B1 into the retina of 4 weeks old rd1 in-vitro-generated Retinal Neuron Like Cells (RNLCs) as cell therapy in the study [32]. These RNLCs were generated from blood monocytes in a two-step culture procedure and shown to be functionally similar to retinal cells including photoreceptors and can rescue vision perception in the rd1 model [32]. We then evaluated whether the adjunct TGF-B1 treatment could improve the survival of cell therapy and hence rescue vision in the rd1mouse model.

## Methods

This investigation was approved by the Institutional Human Ethics Committee (IHEC#100/17) of the National Institute of Immunology (NII), New Delhi. The investigation on mice was approved by the Institutional Animal Ethics Committee (IAEC# 480/18) of NII, New Delhi. All animal experiments were performed in accordance with the guidelines on the regulation of scientific experiments on animals, Ministry of Environment and Forests (Animal Welfare Division), Government of India. All animal experiments and reporting also adhere to the ARRIVE guidelines. This study did not involve a clinical trial.

The CBA/J, rd1 mice were procured from the Jackson Laboratory. The RP mouse model has a Pde6b3’, 5’-cGMP β subunit phosphodiesterase mutation [33] which causes early onset autosomal recessive RP. It displays progressive retinal degeneration from P8[34] and is devoid of rod photoreceptors in the neural retina at P28 (4 weeks). Hence, all experiments were performed on P28, 4 weeks old mice. The animals were housed in the small animal facility at the National Institute of Immunology (NII). They were kept in individually ventilated cages (IVC) and received ad libitum access to acidified autoclaved water and food. They were maintained at 21-23 °C and a 14-h light-10 h dark cycle.

### Intravitreal injection and Aqua-centesis

The animal was anesthetized with ketamine (1mg/10gm) – xylazine (0.1mg/10gm) and the eyes were dilated using 1% tropicamide. Two drops of Paracaine were applied on each eye as a topical analgesic. Both eyes were injected with 0.5ng or 2ng TGF-B1 in 2μl sterile Dulbecco’s Phosphate Buffered Saline (DBPS). For the 2ng dose, a single injection was given while single injection at an interval of 24 hours were given for the 0.5 ng TGF-B1 dose. For intravitreal injection, the bevel 25μl Hamilton syringe with a 33G needle was placed at the scleral-corneal intersection and slight pressure was applied to insert the bevel of the needle. 2μl volume was released and the eye was checked for trauma by looking for signs of haemorrhage or redness. Aqua-centesis was performed for both eyes for 3 days at an interval of 24 hours via trans-corneal injection. The aqueous flush was performed by injecting and re-drawing 5μl sterile PBS. The aqueous sample was used for cytokine analysis

### Isolation of PBMCs from peripheral blood or buffy coat

The PBMCs were isolated by ficoll density centrifugation method in which the blood was diluted with RPMI-1640 media (1:3) and layered over ficoll (density=1.077) in 2:1 ratio followed by centrifugation at 400g for 50 minutes at RT to obtain a buffy layer. The desired number of cells were plated in a 6-well culture plate and left overnight at 37°C and 5% CO2. Detailed protocol in supplementary file P1.

### iTregs generation

PBMCs were isolated from the buffy coat of healthy donors as previously described. The monocytes adhered to the plastic surface, and the supernatant with T cells and non-adherent lymphocytes was used for further processing.

The cells were then stained with anti-CD45RO negative with APC, anti-CD8 with APC, and anti-CD19 flow cytometry antibodies for 40 mins at 4°C. The cells were then washed with PBS and used for sorting naïve T cells.

Negative sorting was performed to acquire the population of CD45RO-CD8-CD19-cells using FACS Aria (BD, Biosciences, USA). The Foxp3 expression was induced upon TGF-B1 and IL-2 supplementation for 5 days [35-37]. One million cells were plated per well of 24 well CD3, CD28 pre-coated plates for 5 days in iTreg media containing 20 μl/mL IL-2 (Prospects Immunotools, Germany) and 5 ng/mL TGF-B1 (Prospects Immunotools, Germany) in 1X RPMI with 10% FBS and 2% antibiotic-antimycotic. Detailed protocol in supplementary file P2 and flow cytometry strategy in S1.

### Immunocytochemistry staining of cells for Flow Cytometry

The cells were incubated in 1:200 dilution of respective conjugated fluorochrome-conjugated antibodies for 40 mins at 4°C. The cells were then washed in PBS by centrifuging at 300g for 10 minutes. The cell pellet was resuspended in 200μl PBS and run in BD FACS Verse flow cytometer.

### Neutralization assay

To evaluate the stability of induced Foxp3 of Treg cells in an inflammatory milieu, a neutralization assay was performed [38]. Wherein, 20,000 iTreg cells were plated in 24 well plates as triplicate, and increasing concentration of TNFα (0, 0.3, 3, 12 ng/ml) was added to the culture media of RPMI with 2% antibiotic-antimycotic for 24 hours. Post 24 hours, immune marker staining for Foxp3 was performed as previously described for flow cytometry analysis.

### Generation of Reconditioned Monocytes (RM) from PBMCs

PBMCs were isolated from the buffy coat of healthy donors as previously described. The non-adherent cells were removed along with the media and the adherent cells were cultured with media composed of IMDM (Himedia, India), 0.5% serum (Himedia, India), (4ng/ml) IL3 (Prospects Immunotools, Germany), (5ng/ml) MCSF (Prospects Immunotools, Germany) along with (140mM) β-mercaptoethanol (Himedia, India) and 1% antibiotic (Himedia, India). The culture was maintained for 6 days with media change every 3 days [39, 40].

### Generation of RNLCs from Reconditioned Monocytes

The reconditioned monocytes were then directly supplied with serum-supplemented RNLCs media and the culture was continued with media change every 3 days until the cells attained the retinal phenotype at day 8. The RNLCs media contained growth factors (20ng/ml) EGF (Prospects Immunotools, Germany), (0.1μg/ml) retinoic acid (Sigma, USA), (20ng/ml) b-FGF (Prospects Immunotools, Germany), (100mM) taurine (Prospects Immunotools, Germany), (20ng/ml) Insulin growth factor IGF-1 (Prospects Immunotools, Germany), Insulin-Transferrin-Selenium (ITS) supplement and 0.5% ESC grade serum (Himedia, India) and 1% antibiotic (Himedia, India) [32]. The RNLCs were characterized for photoreceptor marker recoverin. (Detailed in supplementary file S2.)

### Q tracker dye staining

Q tracker (Invitrogen, Q25011MP) 10nM labeling solution was prepared by mixing 1μl component A and component B of the Q traker kit and the solution was incubated for 5 minutes at room temperature. 0.2ml of fresh complete growth was added to it and vortexed for 30 seconds. 1 million cells were added to the labeling solution and incubated at 37°C for 60 minutes. The cells were washed twice with a complete growth medium by centrifuging at 300g for 10 minutes. The cells were then resuspended in 2μl PBS and used for transplantation.

### Sub-retinal and sub-conjunctival injection of cells

The animals were anesthetized with ketamine (1mg/10gm) – xylazine (0.1mg/10gm) and the eyes were dilated using 1% tropicamide. A few drops of Paracaine were applied on each eye as a topical analgesic. The mouse was placed in lateral recumbence with the eye to be injected facing up. The skin was retracted towards the body causing the eye to protrude. The needle bevel was inserted up into the corneal of the eye at a 45° angle to relieve ocular pressure. A needle with the cell suspension was injected trans-scleral into the sub-retinal region. Eventually, light pressure was applied to the eye to control bleeding. After injection, ophthalmic Tobrex ointment/povidone-iodine (5% solution) was applied along with an antibiotic solution on the mouse eye to prevent infection. Similarly, cells were injected into the sub-conjunctiva of the mouse.

### Estimation of cytokines

To estimate levels of cytokines, the cytokine bead assay (CBA) was performed for IL-10, TGF-B1, TNFα, and MCP1 as per the instruction manual provided by the company (BD Biosciences, USA). The cytokine beads were added to the sample and incubated in the dark at RT for 3 hours. The samples were washed with wash buffer at 300g for 10 minutes and the pellet was then resuspended in 200μl wash buffer. The samples were analyzed in Facs(verse) and FCAP array software was used for the data analysis.

### Transplantation

To achieve a sustained dose range of 0.5ng/ml-2ng/μl of TGF-B1 (Prospects Immunotools, Germany), the iTregs treated group received 20,000 iTreg cells in the sub-conjunctiva of both eyes, and the untreated group received sterile PBS. rd1 mice of P28 age and of any gender were used in the study. The transplantation was performed at P28 and there were 5 animals per group in each study. The animals were euthanized 30 days post transplantation. The RNLCs treated group received 1 million cells per eye in the sub-retinal layer. In the iTregs and RNLCs treated group, RNLCs were transplanted in the sub-retinal layer at day 4 post iTregs transplantation in the sub-conjunctival region. The RNLCs were transplanted on the 4th-day post iTregs transplantation because our earlier experiments indicated a significant decrease in inflammatory cytokines at 72 hours on administering sustained low doses of TGF-B1, resulting in a less hostile milieu for the engraftment of RNLCs. The RNLCs have displayed the highest vision rescue potential at day 20 transplantation post-transplantation. Thus, Electroretinography (ERG) and behavioral studies were compared on day 20. Further, RNLCs were shown to be viable till day 30, hence all molecular studies were undertaken at day 30 post-transplantation.

### Electroretinography

The animals were dark adapted for 1 hour before the procedure. Pupils of dark-adapted mice were dilated with 1% tropicamide after anaesthetization using ketamine (1mg/10gm) – xylazine (0.1 mg/10gm). The gold active wires were placed on the cornea, the ground probe was inserted in the tail, and the reference electrode was subcutaneously placed between the eyes near the cornea. Dark-adapted, full-field scotopic ERG was performed by simulating the retina by 10cds/m^2^ intensity and 25 amplitude readings of ‘A’ and ‘B’ wave were measured (inbuilt algorithm of LabScribe software) (MICRON III rodent imaging system using LabScribe software, Phoenix Laboratory, USA).

### Light/dark latency test

A box sized 21 × 42 x 25 cm was equally divided into two parts by a partition wall with a 5 cm connecting opening. One side of the chamber was brightly illuminated (400 lux) and covered with a white colored background. While the other chamber was kept dark (50 lux) with a black background. The animal was introduced into the light chamber (LC) and observed for five minutes. The total time spent by the animal in each chamber was recorded. The number of transitions between was recorded to observe its exploratory behaviour.

### Enucleation

The eyeballs were micro-dissected for further analysis. Only the retina was micro-dissected from the eyecup and was mechanically disrupted in sterile PBS to obtain single-cell suspension [41]. The suspension was filtered through a 70-micron nylon filter and centrifuged at 300g for 10 minutes. The pellet was resuspended in PBS and stained for flow cytometry studies.

### Statistical analysis

The data was analyzed and plotted with the help of GraphPad Prism 5 software. The statistical significance of the difference between the two groups was calculated by paired test (Prism 5, GraphPad Software Inc.) Parametric test-test and Mann-Whitney test was performed. p values are mentioned and were characterized as p<0.05 (* significant), p<0.01(**), p<0.001(***) and P<0.0001(****).

## Results

### TGF-B1 Therapy

We first assessed the potential of TGF-B1 in neutralizing the ocular inflammation by administering TGF-B1 either as low doses of intravitreal injection every 24 hours (0.5ng/ml) for 3 days (3 injections) or as a single high dose injection (2ng/ml). The intravitreal doses were decided based on the estimated average TGF-B1 levels in wild-type mice (1ng/ml). Thus, sustained low dose (0.5ng) and single high dose (2ng) was utilized. Untreated mice were not injected with any intervention.

At 72 hours post-administration of TGF-B1, a decrease the in the intra-ocular levels of TNFα upon administering was more pronounced compared to the untreated (Figure 1A). MCP1 appeared to be strongly affected by low doses sustained doses TGF-B1. The repeated doses displayed a significant and sustained drop in TNFα (p=0.0007) levels indicating a need for sustained dosage of TGF-B1.

**Figure 1:**
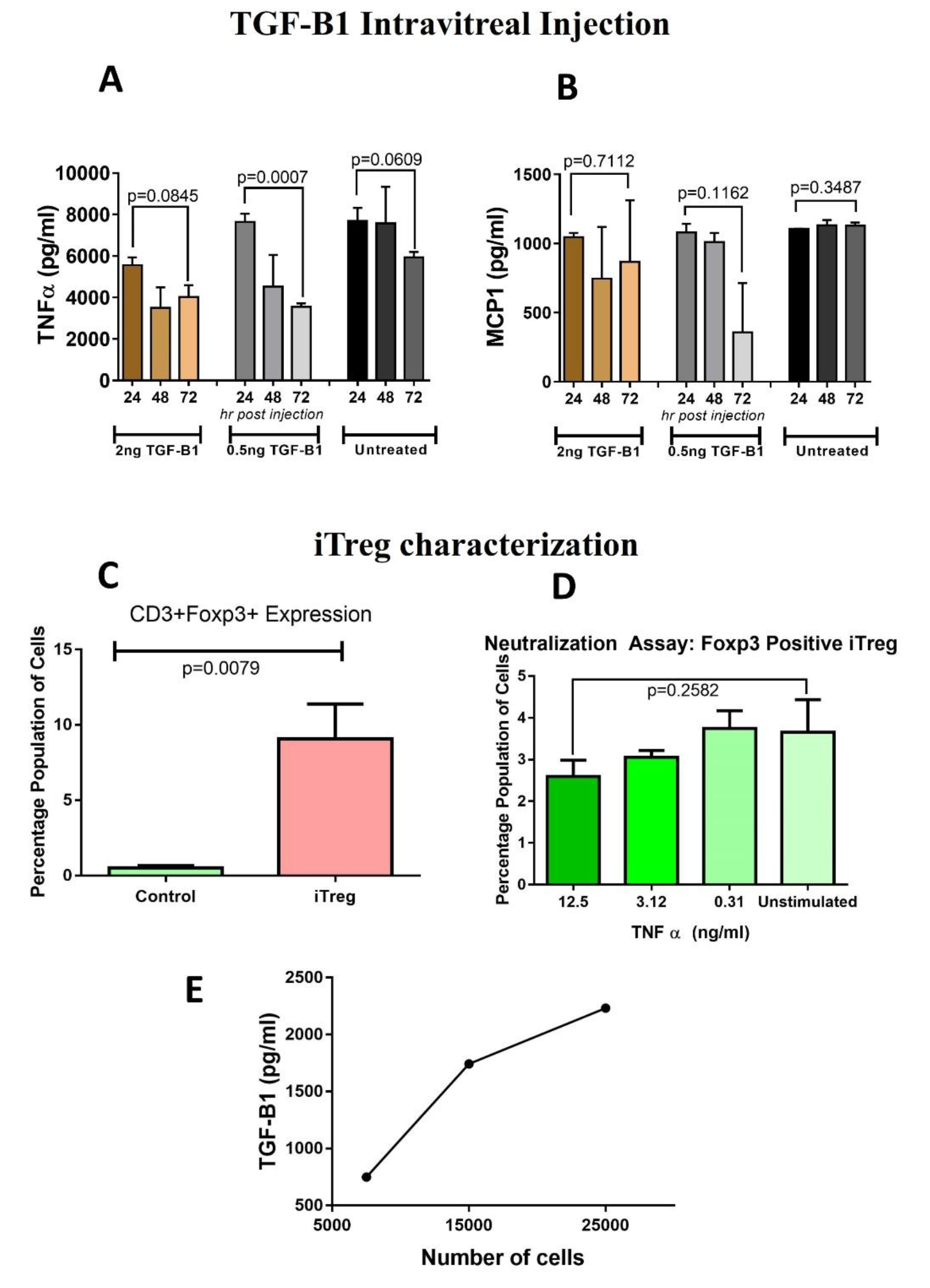
TGF-B1 Intravitreal Injection and iTregs characterization. Panel A: TGF-B1 was administered as multiple low-dose intravitreal injections either every 24 hrs (0.5ng/ml) for 3 days (3 injection total) or as a single high-dose injection (2ng/ml). Aquacentesis was performed every 24 hrs for 3 days and cytokine levels were estimated. The TGF-B1 temporarily reduced the ocular inflammation as seen by a decrease of TNFα and Panel B: MCP1 intra-ocular levels, number of animals per group, n =3. Panel C: On day 5 of the iTregs culture Treg marker Foxp3 was analysed by flow cytometry. iTregs showed a significant increase in Foxp3 expression compared to unstimulated T cells, n=5. Panel D: 20,000 iTreg cells were exposed to increasing concentrations of TNFα (0, 0.3, 3, 12 ng/ml) for 24 hours. The iTreg Foxp3 expression was stable and only slightly suppressed with an increasing TNFα concentration in the media, n=5. Columns show mean and bars over columns are SEM from panel A to D. Panel E: To obtain a maximum of 2ng/ml of TGF-B1, different cell numbers were plated and their cytokine release was estimated; around 20,000 cells released the desired range (0.5ng/ml-2ng/ml) of cytokine concentration, dots show mean and bars over the dots are SEM, n=3.

### iTregs Generation and Characterization

To achieve a sustained low dose of TGF-B1 over a limited time, ex vivo-induced Treg cells (Tregs) were generated from narve human CD3+CD25-T cells.

The resulting iTreg cells expressed a Foxp3 marker significantly higher than the untreated control T cells (P=0.0079) (Figure 1C), and this was stable even in the inflammatory milieu (Figure 1D). To obtain a dose range of 0.5 ng/ml to 2ng/ml of TGF-B1, different numbers of cells were plated and the amount of released TGF-B1 was estimated. It was found that 20,000 cells would release the desired cytokine concentration (Figure 1E). Thus, 20,000 iTregs were transplanted in the sub-conjunctiva of the rd1 mouse in further experiments.

### iTregs and RNLCs transplantation

To achieve a sustained dose of TGF-B1, iTregs and Qtracker-labelled RNLCs were transplanted in the sub-conjunctiva and sub-retinal layers respectively, in different combinations.

We have earlier demonstrated [32] the engraftment of sub-retinally transplanted RNLCs in the rd1 mouse eye by immunohistology and FISH images. In this report we show flow-cytometery data at post 30 days; the single cell suspension of the retina from iTregs transplanted mice displayed a significantly higher number of Qtracker positive RNLCs transplanted in the neural retinal layer than the mice which received only RNLCs (P=0.0143) (Figure 2A) in the flow cytometry study. Flow-cytometery data for analysing transplanted RNLCs is given Supplementary Details, Table ST1.

**Figure 2:**
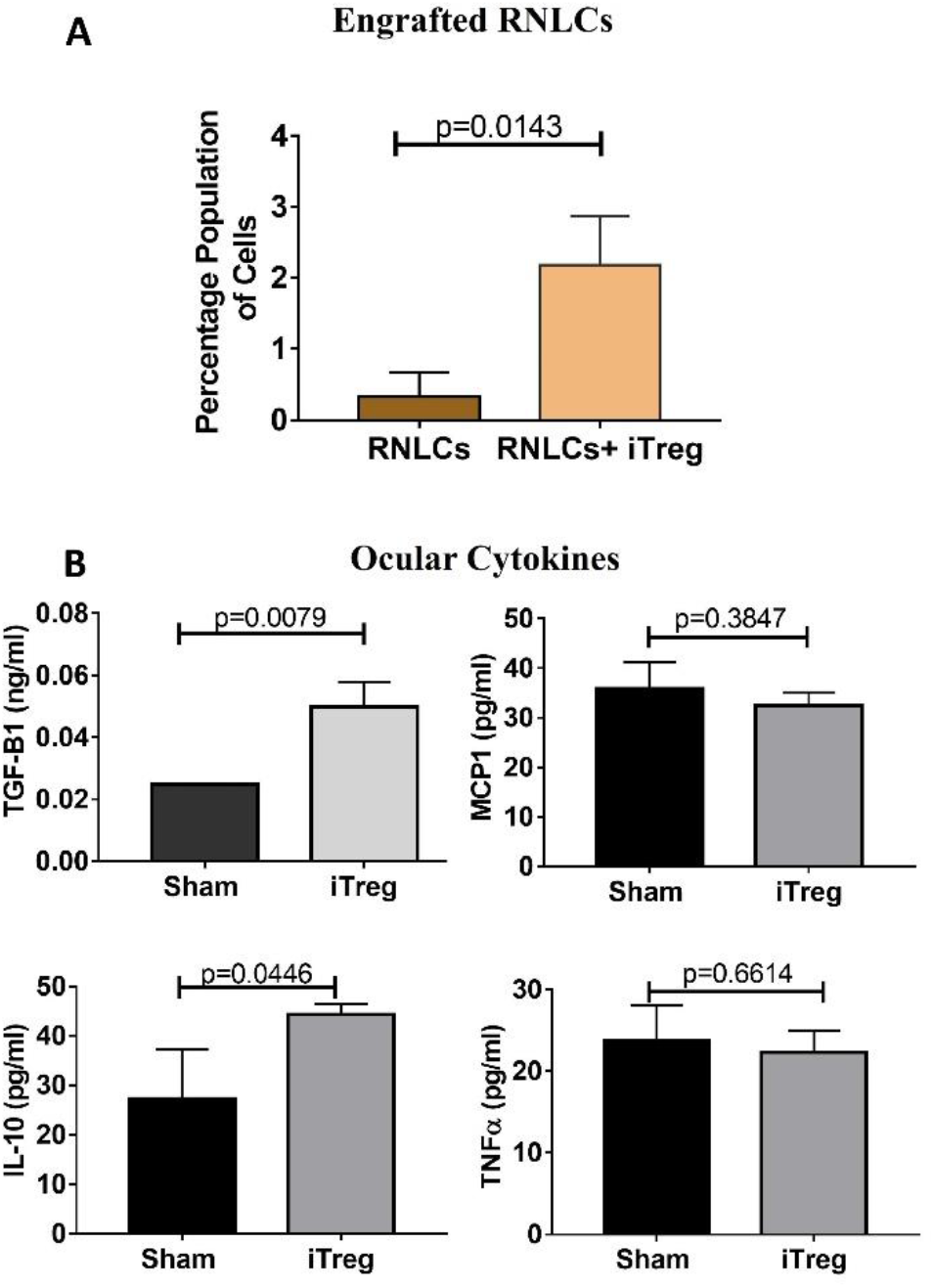
iTregs and RNLCs transplantation. Panel A: The iTregs treated mice displayed a significant increase in the survival of RNLCs transplanted in the retinal layer than the mice which received only RNLCs. Panel B: The ocular cytokine profile at day 30 post-administration indicated a significant increase in anti-inflammatory cytokines TGF-B1 and IL-10. Columns show mean and bars over columns are SEM, n=3 for panel A and B.

At post 30 days post transplantation, there was a significant increase in anti-inflammatory cytokines TGF-B1 (p=0.0079) and IL-10 (p=0.0446) (Figure 2B). Further, a nominal decrease, albeit statistically insignificant, in pro-inflammatory cytokines MCP1 and TNFα was observed (Figure 2B) in the total (aqueous plus vitreous) fluid.

### Post Transplantation Studies

#### Electroretinography

At day 20 post transplantation, animals were subjected to ERG and behavioural studies. The RNLCs post iTregs transplantation/ RNLCs-iTregs mediated vision rescue was significantly higher than only iTregs treated (p=0.0468) and untreated (p=0.0463) animals. The A wave function of ERG indicates photoreceptor function and there was a significant increase in A wave function post iTregs adjunct therapy (Figure 3A). Similarly, the B wave is indicative of secondary neuronal function and there was significant vision conservation upon iTregs adjunct therapy compared to untreated (p=0.03) and only iTregs group (p=0.04).

**Figure 3:**
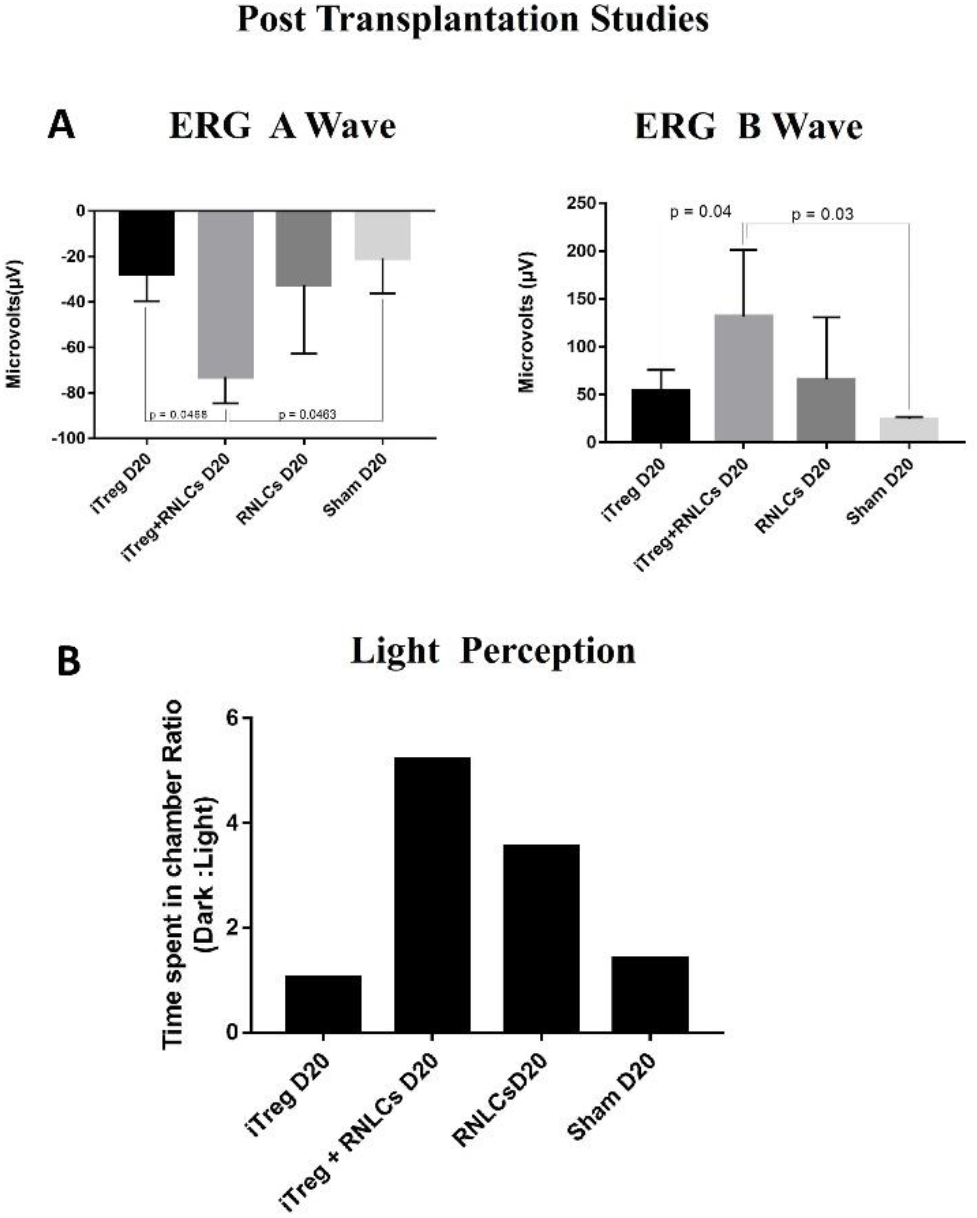
Post Transplantation Studies. Panel A: There was a significant increase in ERG A wave function in the group receiving RNLCs after iTregs treatment at day 20 compared to untreated, n=5. ERG B wave is indicative of secondary neuronal function and there was significant vision conservation upon iTregs therapy. Panel B: The dark latency study evaluated the photophobic response to bright light. The dark:light ratio was highest in the iTregs with RNLCs treated rd1 mouse. Columns show mean and bars over columns are SEM, n=5.

### Behavioural Study

Figure 3B shows the ratio of time spent by the mouse in the dark and light chambers among different groups at day 20 post transplantation. This ratio was highest in the group which received RNLCs post iTreg therapy. It was followed by only the RNLCs group.

## Discussion

The study demonstrates that TGF-B1 therapy can improve the functional engraftment of cell therapy in the rd1 mouse model, despite the ocular inflammation caused by retinal degeneration. This finding suggests that TGF-B1 may help create a more favourable environment for cell-based interventions.[8, 12-15]. Thus, Treatment with TGF-B1 has the potential to lower inflammation and boost the effectiveness of cell-based therapies.

Our study examined the impact of TGF-B1 on inflammatory cytokines, particularly TNFα and MCP1, to gauge its effectiveness in reducing ocular inflammation. TNFα has been shown to exacerbate damage to the blood retinal barrier, impeding the secretion of anti-inflammatory agents.[10, 14, 23, 42, 43]. The MCP1 causes detrimental microglia activation and attracts macrophages into the eye leading to ocular inflammation[10, 14, 23, 24, 43, 44].

The experiments indicate that supplementing external TGF-B1 can effectively decrease inflammatory cytokine levels in the eye. However, to sustain this reduced inflammation, a continuous release of TGF-B1 is imperative. However, administering multiple corneal injections can lead to retinal bleeding, which can be deleterious.[45]. Additionally, a high dose of extrinsic of TGF-B1 over a long period can cause severe changes in intra-ocular pressure and development of cataracts due fibrotic response[46].

Thus, we attempted to modulate the ocular inflammatory conditions using low and sustained doses of TGF-B1 from iTregs. The iTregs have been successfully used in corneal transplants for managing anterior chamber inflammation [28]. We generated iTregs from naïve T cells and confirmed the Foxp3 expression. The Foxp3 was found to be stable in inflammatory conditions in the iTregs. Hence, the requisite number of iTregs was estimated and transplanted in the sub-conjunctiva.

The transplantation of 20,000 iTregs was intended to facilitate the secretion of cytokines which could release a required dose of up to 2ng/ml of anti-inflammatory cytokines into the eye [28]. In addition, the presence of TGF-B1 is expected to inhibit the invasion of the peripheral immune system into the conjunctiva [37].

Previous studies have demonstrated that *in vitro* [35-37], iTregs remain viable for up to 7 days. Therefore, sub-conjunctival transplantation of iTregs could potentially prevent the entry of cellular debris into the retinal area through their clearance via the blood network. It is challenging to observe the transplantation of iTregs due to the presence of resident lymphocytes. As a result, molecular, behavioural, and functional evaluations were conducted.

To assess the potential of using iTregs in cell-based therapy research, we utilized RNLCs that were generated *in-vitro* and have the ability to integrate into retina of rd1 mice [32]. Transplantation of iTregs offers an added advantage of continuous release of anti-inflammatory cytokines that can effectively curb inflammation during the transplantation procedure. Moreover, the implantation of RNLCs in the eye has been proven to trigger anti-inflammatory responses, as reported by Mishra et al. [32].

At 30 days post transplantation, the levels of TGF-B1 and IL-10, which are known to reduce inflammation, were significantly increased. On the other hand, the levels of TNFα and MCP1, which are inflammatory cytokines, were remarkably reduced. Moreover, a notable decrease in cellular infiltrates was observed, as illustrated in the Supplementary Details, Figure. S3 The anti-inflammatory ocular environment created by this shift in cytokine levels contributed significantly to the engraftment and survival of cell therapy in the retina. There was a significant increase in the number of transplanted RNLCs in the rd1 retina, compared to the RNLCs treated group. This increase in RNLCs may have contributed to the preservation of vision, particularly in the B wave of the ERG. The dark latency test confirmed this, as the dark:light ratio was highest in the iTregs and RNLCs treated rd1 mouse at day 20, i.e. the mouse spent more time in the dark chamber.In contrast, the untreated mouse did not display typical photophobic behaviour.

Overall, these findings suggest that the anti-inflammatory ocular environment created by the increased levels of TGF-B1 and IL-10 played a crucial role in promoting better engraftment and survival of transplanted RNLCs, which in turn contributed to the preservation of vision in the rd1 mice.

## Conclusions

This pilot study offers a novel perspective on potential anti-inflammatory therapies for RP by exploring the effectiveness of using TGF-B1 secreting iTregs to mitigate inflammation caused by retinal degeneration. The study suggests that this approach may improve the outcome of cell therapy for RP.

## Data Availability

All data produced in the present work are contained in the manuscript

## Acknowledgments

Electroretinography recordings were made at the Ocular Pharmacology and Pharmacy Division, Dr. RP Centre, All India Institute of Medical Sciences, New Delhi. We are grateful to Professor Velpandian Thirumurthy for the same.

## Funding

This work was supported by a grant received from the Department of Science and Technology, Government of India (DST/ICPS/EDA/2018) and the core grant received from the Department of Biotechnology, Government of India to the National Institute of Immunology, New Delhi. P.S. was granted a research fellowship by the Department of Biotechnology.

## Competing interests

The authors declare that they have no competing interests.

## List of abbreviations

RP: Retinitis Pigmentosa
TGF-B1: Transforming Growth Factor-Beta1
RNLCs: Retinal Neuron Like Cells
iTregs: induced-Treg cells
TNFα: Tumor necrosis factor-alpha
AAV: Adeno-Associated Virus
GMP: Guanosine 3’,5’-cyclic monophosphate cyclic
IVC: Individual Ventilated Cages
CBA: Cytokine Bead Assay
ERG: Electroretinography
MCP1: Monocyte Chemoattractant Protein-1

## Notes

### Competing Interest Statement

The authors have declared no competing interest.

### Author Declarations

This investigation was approved by the Institutional Human Ethics Committee (IHEC#100/17) of the National Institute of Immunology (NII), New Delhi. The investigation on mice was approved by the Institutional Animal Ethics Committee (IAEC# 480/18) of NII, New Delhi.

