## supplementary file for "iTreg mediated TGF-Β1 therapy improves functional engraftment of cell therapy in rd1 Retinitis Pigmentosa mouse model"

#### *Contents*

**Protocol details**
P1. Isolation of Peripheral Blood Mononuclear Cells (PBMCs) page 2
P2. iTreg Generation page 2

**Additional data**
S1. Gating Strategy for negative sorting page 3
S2. RNLC characterization page 4
S3. Cellular infiltration page 5
ST1. Flow-cytometery data for analysing transplanted RNLCs page 6

### 1. Isolation of PBMCs from peripheral blood or buffy coat

The PBMCs was isolated by ficoll density centrifugation method in which the blood was diluted with RPMI-1640 media (1:3) and layered over ficoll (density=1.077) in 2:1 ratio followed by centrifugation at 400g for 50 minutes at RT to obtain a buffy layer. The layer was collected and diluted with PBS and centrifuged at 300g for 10 minutes at room temperature to obtain a pellet of PBMCs. The pellet was further re-suspended in complete IMDM and plated in culture plate. The viable cells are counted by trypan blue exclusion method using haemocytometer and desired number of cells are plated in 6 well culture plate and left overnight in 37$^{0}$C and 5% $CO_2$ for adherence.

### 2. iTreg generation

PBMCs were isolated from buffy coat of healthy donors as previously described. The PBMCs were cultured in T75 flask with 10% FBS 2% antibiotic-antimycotic in complete RPMI media, overnight at 37◦C. The monocytes adhered to the plastic surface, therefore the supernatant with T cells and non-adherent lymphocytes was used for further processing. The supernatant was then centrifuged at 300g at 4◦c for 10mins. The pellet was washed with PBS and the cells were counted. The cells were then stained with anti-CD45RO negative with APC, anti-CD8 with APC and anti-CD19 with or 40mins at 4◦c. Thereafter, the cells were washed with PBS to remove unbound antibodies and resuspended in 20%FBS 3% antibiotic-antimycotic in complete 1X RPMI at the dilution of 10 million cells/ml. Cells were then passed through a 40µm cell strainer. Unstained and single colour control were also prepared for voltage settings and compensation. Negative sorting was performed to acquire the population of CD45R0- CD8- CD19- cells for naïve T cells gating on scatter plot of lymphocytes using FACS Aria (BD, Biosciences, USA) (Gating strategy is given in supplementary details, S1). The sorted cells were collected in tubes containing RPMI with 10 % FBS and 2% antibiotic-antimycotic. The cells were centrifuged at 300g for 10 mins. The pellet was resuspended in iTreg media containing IL-2 (20 µl/mL) and TGF-B1 (5 ng/mL) in 1X RPMI with 10% FBS and 2% antibiotic-antimycotic. One million cells were plated per well of 24 well CD3, CD28 pre-coated plates for 5 days.

### S1. Negative Sorting Gating Strategy

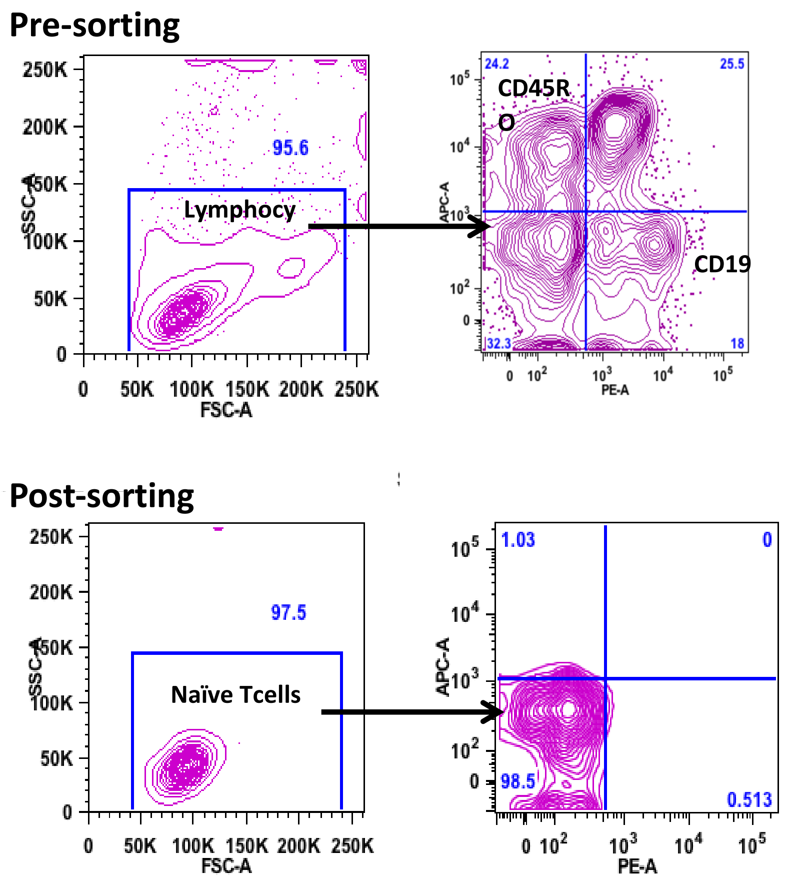


Figure S1: Negative sorting was performed to acquire the population of CD45R0- CD8- CD19- by the cells gating on scatter plot of lymphocytes using FACS Aria (BD, Biosciences, USA). One million cells were plated per well of 24 well CD3, CD28 pre-coated plates for 5 days in iTreg media containing IL-2 (20 U/mL) and TGF-B1 (5 ng/mL) in 1X RPMI with 10% FBS and 2% antibiotic.

### S2. RNLC Characterization

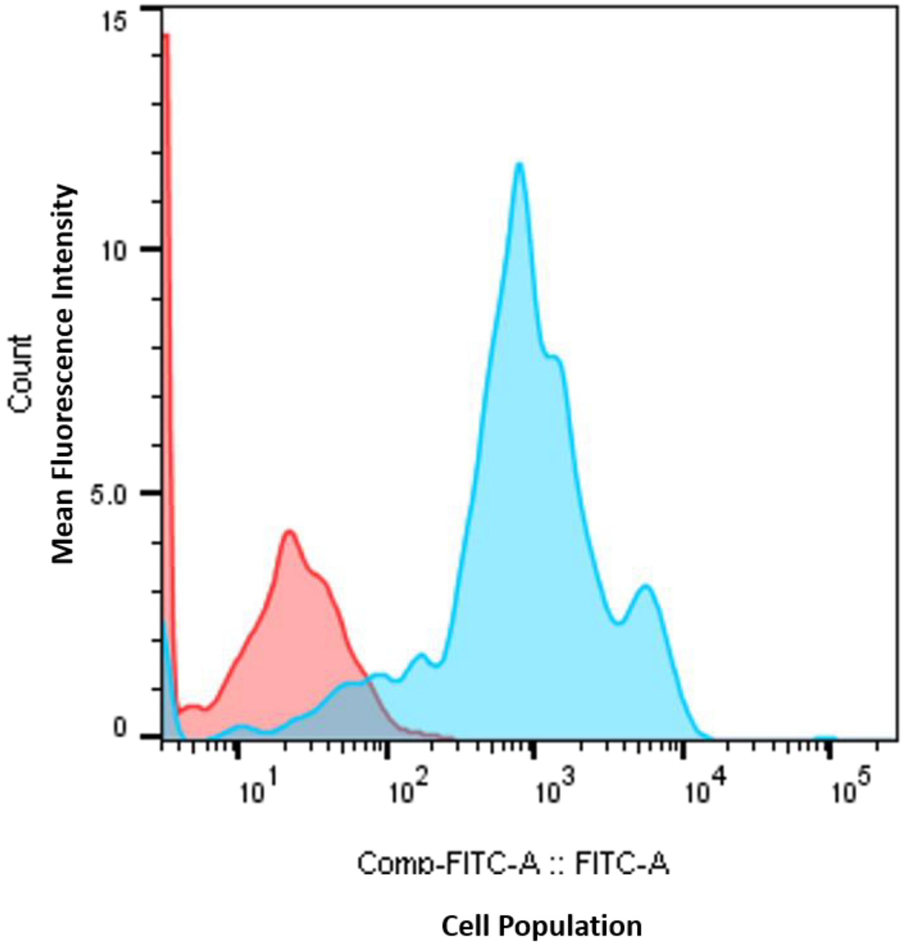


Figure S2: RNLC generation and characterization. At the end of 14 days culture the cells were checked for the expression of photoreceptor marker. The RNLCs expressed recoverin (FITC) in the facs study and were used for transplantation n=5.

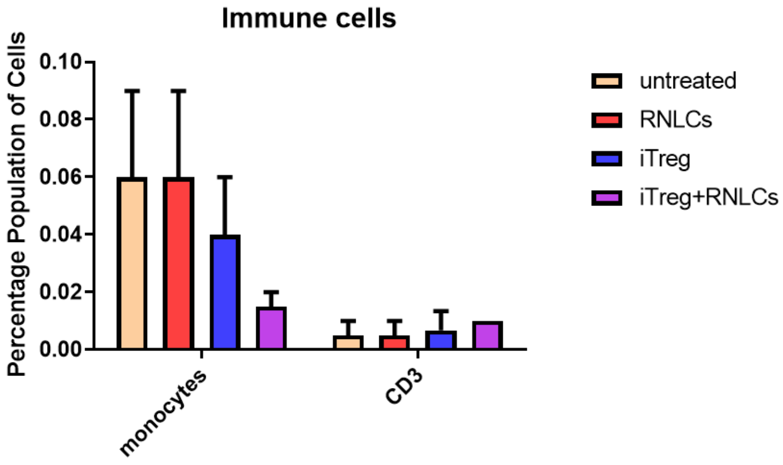


Figure S3: Cellular infiltration. There was no significant increase in the cellular infiltration post-transplantation of iTregs and RNLCs. The retina was dissected and the single cell suspension was studied for immune cell response of monocytes CD14 and T cell Cd3 by flow cytometry at 30 days post-transplantation.

Table ST1: Flow-cytometery data for analysing transplanted RNLCs

| | | |
|---|---|---|
| **RNLCs** | | |
| r1.fcs | | 10000 |
| retinal cells | 81.2 | 8118 |
| Fitc | 0.73 | 59 |
| **RNLCs** | | |
| r2.fcs | | 10000 |
| retinal cells | 83.7 | 8368 |
| fitc | 0.12 | 10 |
| **RNLCs** | | |
| r3.fcs | | 10000 |
| retinal cells | 84.6 | 8462 |
| fitc | 0.13 | 11 |
| **RNLCs+iTreg** | | |
| rt1.fcs | | 10000 |
| retinal cells | 88.4 | 8836 |
| fitc | 2.18 | 193 |
| **RNLCs+iTreg** | | |
| rt2.fcs | | 10000 |
| retinal cells | 81.3 | 8129 |
| fitc | 2.87 | 233 |
| **RNLCs+iTreg** | | |
| rt3.fcs | | 10000 |
| retinal cells | 81.3 | 8131 |
| fitc | 1.49 | 121 |

Table ST1: The table describes the number of Qtracker positive cells estimated per 10000 retinal cells in the flow cytometry study. The numbers are indicative of the survival of the cell therapy at day 30 post-transplantation of RNLCs in the sub-retinal layer of the mice retina.
